# Cognitive behavioral therapy for insomnia as a suicide prevention strategy: a protocol for a systematic review and meta-analysis

**DOI:** 10.1101/2025.06.29.25330506

**Authors:** Cagdas Türkmen, Carlotta L. Schneider, Yuki Furukawa, Jens H. van Dalfsen, William V. McCall, Wilfred R. Pigeon, Andrew S. Tubbs, Michael L. Perlis, Dieter Riemann, Kai Spiegelhalder, Knut Langsrud, Håvard Kallestad, Elisabeth Hertenstein

## Abstract

**Background:** Insomnia is a highly prevalent and debilitating sleep-wake disorder, with growing evidence indicating that it is an independent risk factor for suicidal ideation and behaviors. Cognitive behavioral therapy for insomnia (CBT-I) is the recommended first-line treatment for chronic insomnia. However, its effect on suicidal ideation and behaviors in those with insomnia has not been well-characterized. Thus, the aim of the planned meta-analysis is to quantify the effects of CBT-I on suicidal ideation, suicidal behaviors and suicide deaths in adults with insomnia with and without comorbidities.

**Methods:** The planned study will include randomized controlled trials (RCTs) comparing CBT-I with a control condition with no presumed strong effect on insomnia in individuals with insomnia according to standardized diagnostic criteria or a clinically relevant screening score. Both published and unpublished RCTs will be retrieved through a systematic search in major databases and trial registries. The primary outcomes include 1) suicidal ideation, 2) suicidal behaviors, and 3) suicide deaths, assessed post-treatment and at follow-ups. We will only consider RCTs reporting suicide-related outcomes and/or enrolling participants with suicidal ideation or behaviors. For continuous data, a random-effects meta-analysis will be conducted to estimate (standardized) mean differences. In the case of categorical data, a random-effects logistic regression meta-analysis model will be used. The risk of bias of the primary outcomes will be evaluated using the Cochrane Risk of Bias 2 tool. The certainty of evidence will be assessed using GRADE. All analyses will be conducted in the R software.

**Discussion:** The planned meta-analysis will provide a synthesis of the effects of CBT-I on suicide-related outcomes in individuals with insomnia. The findings could have important implications for integrating sleep-focused interventions into suicide prevention strategies and inform clinical practice, particularly for individuals with comorbid insomnia and increased suicide risk.

**Systematic review registration:** PROSPERO-ID CRD420250628820

**Review question:** How effective is cognitive behavioral therapy for insomnia (CBT-I) in reducing suicidal ideation, suicidal behaviors and suicide deaths among adults with insomnia with and without comorbidities?

## Background

Insomnia is a highly prevalent and debilitating sleep-wake disorder, affecting approximately 1 in 10 individuals in the general population (1). It is associated with reduced health-related quality of life (2), impaired work productivity (3) and an increased risk of developing psychiatric disorders (4). Accumulating evidence indicates that insomnia is also a significant risk factor for suicidal ideation and behaviors across various age groups (5–10). Thus, the effective treatment of insomnia is of high public health importance.

Cognitive behavioral therapy for insomnia (CBT-I) is the recommended first-line treatment for chronic insomnia among adults (11). Recent meta-analyses have shown that CBT-I is effective for the treatment of insomnia in individuals with comorbid mental health and alcohol use disorders (12–14), where insomnia can exacerbate symptoms and increase the risk of relapse (15, 16). While CBT-I may reduce comorbid symptom severity (12), there is a paucity of evidence synthesis addressing whether CBT-I may also reduce suicidal ideation and behaviors.

Since 2010, there has been a call to action to evaluate sleep interventions as a strategy for suicide prevention (17). There has been a slow but steady response to this call, with initial observational studies suggesting that CBT-I may reduce suicidal ideation (18, 19). In recent years, CBT-I has gained momentum through several randomized controlled trials contributing to the evidence base (20–24). Although the latest meta-analysis reported a modest effect of sleep interventions (including behavioral and pharmacological treatments) in reducing suicidal ideation, the findings were limited by substantial variability among the relatively few included studies (25). The planned meta-analysis aims to expand upon this work by updating the evidence with newly published studies, incorporating unpublished data, and narrowing the focus to CBT-I. Specifically, we aim to quantify the effects of CBT-I, compared with control conditions, on suicidal ideation, suicidal behaviors and suicide deaths among adults with insomnia with and without comorbidities.

## Methods

The information in this protocol is reported in accordance with the guidelines of the Preferred Reporting Items for Systematic Review and Meta-Analysis Protocols (PRISMA-P) (26, 27). The completed PRISMA-P checklist is provided in **Supplement 1**. The protocol was prospectively registered on PROSPERO (CRD420250628820).

### Study selection and selection criteria

Two reviewers will independently screen the titles and abstracts of all studies, and will retrieve and review full-text reports of potentially relevant studies to determine their eligibility. We will include published and unpublished records reporting on randomized controlled trials (RCTs) that compare CBT-I in any delivery format to control conditions with no presumed strong effect on insomnia (i.e., placebo, sleep hygiene, self-help, waitlist, no treatment) in parallel-group designs. CBT-I must include stimulus control or sleep restriction as behavioral components, in accordance with recent network meta-analyses identifying these as critical components (28, 29). CBT-I may be combined with other treatments (e.g., specific to a psychiatric disorder), provided that the control group receives the same treatment. We will consider adults (≥ 18 years) of any gender with insomnia with or without comorbidity according to diagnostic criteria (i.e., the Diagnostic and Statistical Manual of Mental Disorders (30, 31), the International Classification of Sleep Disorders (32) or Diseases (33, 34)), or a validated, clinically meaningful screening score. We will only consider studies reporting suicide-related outcomes and/or enrolling participants with suicidal ideation or behaviors. Disagreements regarding study selection will be resolved by consensus or, if necessary, with the involvement of another member of the review team.

### Search strategy

The following databases and registers were systematically searched to identify records published from inception through 21/03/2025: Medline, Embase, Cochrane Library, PsycInfo and ClinicalTrials.gov. No language restrictions were applied. An RCT filter was applied to all databases except the Cochrane Library. The search will be complemented by screening the reference lists of the included studies and of relevant systematic reviews on this topic (25, 35). Additionally, forward citation searches (i.e., identifying studies that have cited the included studies and relevant systematic reviews on this topic) will be performed to capture any further eligible records. Search strings and the number of hits for each database/register are provided in **Supplement 2**.

### Primary and secondary outcomes

The primary outcomes include suicidal ideation, suicidal behaviors and suicide deaths, assessed post-treatment and at follow-up(s). The secondary outcomes include insomnia severity, also measured post-treatment and at follow-up(s), as well as depression severity and hopelessness which are key correlates of suicidal ideation, behaviors and suicide deaths (36). Based on preliminary searches, both the primary and secondary outcomes (with the exception of suicide deaths) are commonly measured using continuous outcome measures. Thus, continuous data will be prioritized over categorical data in the analyses.

### Record management

Records will first be imported into EndNote (Clarivate, Version 21, 2023) (37) and deduplicated using the software’s built-in functions. The deduplicated records will then be exported from EndNote to an Excel spreadsheet for formal screening. Records will initially be coded as clearly irrelevant (excluded prior to full-text review based on title and/or abstract) or potentially relevant (proceeding to full-text review). Following full-text review, potentially relevant records will be coded as either included or excluded, with reasons documented for each exclusion.

### Data extraction

Two reviewers will independently extract data from all included trials for the following variables: authors, year of publication, number of participants in the CBT-I/control groups at each timepoint, age of participants at baseline, percentage of women at baseline, race/ethnicity, number of follow-up assessments, time between baseline and each follow-up, criteria for insomnia, outcome measures, CBT-I components, number of CBT-I sessions, treatment duration, and details regarding control conditions. This information will be summarized narratively in the text and/or presented in tabular form.

Outcome data will be extracted independently by two reviewers using a pre-defined Excel sheet for the meta-analyses. Depending on the frequency of continuous versus categorical data reporting, we may opt to harmonize the data to enhance statistical power and reduce the number of analyses by reaching out to study authors. For instance, if most studies reported continuous data and it is feasible to obtain continuous data from studies that reported categorical data (e.g., those that transformed continuous data into binary outcomes using cut-off scores), we will request these data from the principal investigators.

Principal investigators of both unpublished and published studies will be contacted up to three times, with a two-week interval between each attempt, to request unpublished data or address other relevant inquiries.

### Statistical analysis

For pairwise comparisons informed by ≥ 3 RCTs, we will perform a meta-analysis if the RCTs are sufficiently homogeneous with respect to design and comparator. For continuous data, mean differences (MDs) or standardized mean differences (SMDs), along with 95% confidence intervals (CIs), between the CBT-I group versus control groups will be calculated for the outcomes post-treatment and at follow-up(s). MDs will be used when all studies measured an outcome on the same scale, while SMDs will be used when multiple scales are used across studies. In the case of SMDs, Hedges’ g will be used as a measure of effect size. We will rely on post- and (follow-up-) means, instead of a change score between baseline and post-(and follow-up-) means. Based on the assumption that there is a distribution of true effect sizes rather than a single true effect size, a random-effects model will be used when pooling the primary studies, in line with the recommendation by Borenstein et al. (2010) (38). Forest plots will be presented to visualize the results. The degree of heterogeneity between studies will be assessed using I^2^, which describes the proportion of total variation in estimated effect sizes attributable to differences among the studies. An I^2^ value of 50% or higher is commonly considered an indicator of heterogeneity. As an additional measure of heterogeneity and to estimate the range of effect sizes in future studies, corresponding 95% prediction intervals will be calculated (39, 40).

For categorical data (yes/no), odds ratios (ORs) and 95% CIs will be estimated using a random-effects logistic regression model. Suicide-related outcomes, particularly suicide deaths, are expected to be rare (i.e., low counts). For rare events, the Mantel-Haenszel method will be used, which avoids continuity corrections that might bias results (41, 42). To evaluate the robustness of the results, we will compare the results of the inverse variance model (which assumes a common treatment effect) and those of the Mantel-Haenszel method. If notable discrepancies arise between the methods, only the Mantel-Haenszel method will be used. Forest plots will be used for the visualization of the results, applying a 0.5 continuity correction for studies with zero events in one treatment arm. We will report the I^2^ statistic, along with its 95% CI, as an indicator of heterogeneity for all analyses. Prediction intervals will be calculated as an additional indicator for the degree of heterogeneity and to estimate the effect size range in future studies (39, 40).

All analyses will be conducted in R using the meta package (43). Although we anticipate conducting meta-analyses for the primary and secondary outcomes, we will provide a systematic narrative synthesis if quantitative synthesis is not appropriate (e.g., due to insufficient data). Information will be presented in the text and in tabular form to summarize and explain the characteristics and findings of the included studies. The narrative synthesis will assess the relationships and results within and between the included studies

### Subgroup analyses

Assuming that sufficient data are available, meta-regression will be used to explore potential subgroup differences based on study characteristics, including age group, psychiatric comorbidities, CBT-I delivery format (e.g., in-person vs. digital) and treatment setting (e.g., outpatient vs. inpatient).

### Sensitivity analyses

To assess the robustness of all synthesized results, we will conduct sensitivity analyses excluding studies at high overall risk of bias.

### Publication bias

Provided that ≥ 10 studies are included (44), the presence of potential publication bias (or ‘small-study effects’) will be assessed by examining asymmetry in a contour-enhanced funnel plot (45) using the primary outcomes.

### Risk of bias

Two reviewers will independently assess the risk of bias of the primary outcomes in each study using the Cochrane Risk of Bias 2 (RoB 2) tool (46). The assessment will cover the following domains:

1. bias arising from the randomization process,
2. bias due to deviations from the intended interventions,
3. bias due to missing outcome data,
4. bias in measurement of the outcome, and
5. bias in selection of the reported result.

Each domain will be rated as “low risk of bias”, “some concerns”, or “high risk of bias”. The overall risk of bias of each primary outcome will be determined by the least favorable rating among the domains. The assessments will be managed using the RoB 2 Excel tool. Disagreements will be resolved by consensus or, if necessary, with the involvement of another member of the review team.

### Certainty of evidence

Two reviewers will independently assess the certainty of the evidence for the primary outcomes using the Grading of Recommendations, Assessment, Development and Evaluation (GRADE) approach (47). Based on GRADE guidelines (48), the following factors will be considered: 1) risk of bias (study limitations), 2) inconsistency of results (heterogeneity in the meta-analysis), 3) indirectness (relevance to the research question), 4) imprecision (small sample sizes, wide CIs), and 5) publication bias. The certainty in the body of evidence will be rated as high, moderate, low or very low. Justifications will be provided for decisions to downgrade or upgrade the certainty of the evidence, as well as for the importance rating of each outcome. Disagreements will be resolved by consensus or, if necessary, with the involvement of another member of the review team.

## Discussion

The planned meta-analysis will offer a synthesis of the current evidence on the effects of CBT-I on suicide-related outcomes in individuals with insomnia. Given the strong and growing evidence linking insomnia with suicidal ideation and behaviors (5–10), the findings could have important implications for clinical practice and suicide prevention strategies. Notably, CBT-I in digitally delivered formats (digital CBT-I) offers a scalable, low-cost, and accessible alternative to face-to-face CBT-I (49), making it a promising component of broader public health approaches to suicide prevention.

However, it is important to acknowledge that RCTs in this specific area of research have only recently begun to emerge (20), and the current evidence base may still be limited in both size and scope. As such, the planned meta-analysis may also play a critical role in identifying gaps in the literature, highlighting important methodological considerations, and informing the design of future studies. Beyond its clinical relevance, the review could serve as a foundation for advancing research into the mechanisms underlying the relationship between insomnia and suicidal ideation/behaviors, ultimately contributing to the development of more targeted prevention strategies.

## Data Availability

Not applicable.

## Abbreviations

CBT-I: Cognitive behavioral therapy for insomnia
CI: Confidence interval
GRADE: Grading of Recommendations, Assessment, Development and Evaluation
PRISMA-P: Preferred reporting items for systematic review and meta-analysis protocols
PROSPERO: International prospective register of systematic reviews
RCT: Randomized controlled trial

## DECLARATIONS

### Ethics approval and consent to participate

The planned review does not require ethical approval or consent to participate.

### Consent for publication

The planned review will not incorporate any individual person’s data in any form. Thus, consent for publication is not required. The findings will be published in a peer-reviewed journal and may be presented at international conferences.

### Availability of data and materials

Not applicable.

### Authors’ contributions

CT, the guarantor of the review, conceived the study and wrote the first draft of the protocol. All authors contributed to the study design and provided intellectual input on the first draft. All authors approved the final version of the protocol to be published.

## Acknowledgements

We thank the librarian Dipl.-Bibl. Volker Braun from the Library of the Medical Faculty Mannheim, University of Heidelberg, for his assistance in developing the systematic search strategy and managing the records. We also extend our gratitude to the Central Institute of Mental Health in Mannheim, Germany, for supporting this work through institutional open access funding.

## References

1. Morin CM, Jarrin DC. Epidemiology of Insomnia: Prevalence, Course, Risk Factors, and Public Health Burden. Sleep Med Clin. 2022;17(2):173–91.

2. Kyle SD, Morgan K, Espie CA. Insomnia and health-related quality of life. Sleep Med Rev. 2010;14(1):69–82.

3. Chalet FX, Albanese E, Egea Santaolalla C, Ellis JG, Ferini-Strambi L, Heidbreder A, et al. Epidemiology and burden of chronic insomnia disorder in Europe: an analysis of the 2020 National Health and Wellness Survey. J Med Econ. 2024;27(1):1308–19.

4. Hertenstein E, Feige B, Gmeiner T, Kienzler C, Spiegelhalder K, Johann A, et al. Insomnia as a predictor of mental disorders: A systematic review and meta-analysis. Sleep Med Rev. 2019;43:96–105.

5. Pigeon WR, Pinquart M, Conner K. Meta-analysis of sleep disturbance and suicidal thoughts and behaviors. J Clin Psychiatry. 2012;73(9):e1160–7.

6. Bernert RA, Kim JS, Iwata NG, Perlis ML. Sleep disturbances as an evidence-based suicide risk factor. Curr Psychiatry Rep. 2015;17(3):554.

7. Baldini V, Gnazzo M, Rapelli G, Marchi M, Pingani L, Ferrari S, et al. Association between sleep disturbances and suicidal behavior in adolescents: a systematic review and meta-analysis. Front Psychiatry. 2024;15:1341686.

8. Liu RT, Steele SJ, Hamilton JL, Do QBP, Furbish K, Burke TA, et al. Sleep and suicide: A systematic review and meta-analysis of longitudinal studies. Clinical Psychology Review. 2020;81:101895.

9. Malik S, Kanwar A, Sim LA, Prokop LJ, Wang Z, Benkhadra K, et al. The association between sleep disturbances and suicidal behaviors in patients with psychiatric diagnoses: a systematic review and meta-analysis. Systematic Reviews. 2014;3(1):18.

10. Liu J-W, Tu Y-K, Lai Y-F, Lee H-C, Tsai P-S, Chen T-J, et al. Associations between sleep disturbances and suicidal ideation, plans, and attempts in adolescents: a systematic review and meta-analysis. Sleep. 2019;42(6).

11. Riemann D, Espie CA, Altena E, Arnardottir ES, Baglioni C, Bassetti CLA, et al. The European Insomnia Guideline: An update on the diagnosis and treatment of insomnia 2023. Journal of Sleep Research. 2023;32(6):e14035.

12. Hertenstein E, Trinca E, Wunderlin M, Schneider CL, Züst MA, Fehér KD, et al. Cognitive behavioral therapy for insomnia in patients with mental disorders and comorbid insomnia: A systematic review and meta-analysis. Sleep Med Rev. 2022;62:101597.

13. Türkmen C, Schneider CL, Viechtbauer W, Bolstad I, Chakravorty S, Miller MB, et al. Cognitive behavioral therapy for insomnia across the spectrum of alcohol use disorder: A systematic review and meta-analysis. Sleep Medicine Reviews. 2025;80:102049.

14. Furukawa Y, Nagaoka D, Sato S, Toyomoto R, Takashina HN, Kobayashi K, et al. Cognitive behavioral therapy for insomnia to treat major depressive disorder with comorbid insomnia: A systematic review and meta-analysis. J Affect Disord. 2024;367:359–66.

15. Brower KJ. Insomnia, alcoholism and relapse. Sleep Med Rev. 2003;7(6):523–39.

16. Khurshid KA. Comorbid Insomnia and Psychiatric Disorders: An Update. Innov Clin Neurosci. 2018;15(3-4):28–32.

17. Pigeon WR, Caine ED. Insomnia and the risk for suicide: Does sleep medicine have interventions that can make a difference? Sleep Medicine. 2010;11(9):816–7.

18. Trockel M, Karlin BE, Taylor CB, Brown GK, Manber R. Effects of cognitive behavioral therapy for insomnia on suicidal ideation in veterans. Sleep. 2015;38(2):259–65.

19. Jernelöv S, Forsell E, Kaldo V, Blom K. Initial Low Levels of Suicidal Ideation Still Improve After Cognitive Behavioral Therapy for Insomnia in Regular Psychiatric Care. Front Psychiatry. 2021;12:676962.

20. McCall WV. Targeting insomnia symptoms as a path to reduction of suicide risk: the role of cognitive behavioral therapy for insomnia (CBT-I). Sleep. 2022;45(12).

21. Pigeon WR, Funderburk JS, Cross W, Bishop TM, Crean HF. Brief CBT for insomnia delivered in primary care to patients endorsing suicidal ideation: a proof-of-concept randomized clinical trial. Transl Behav Med. 2019;9(6):1169–77.

22. Pigeon W, Crean H, Bishop T, Cross W, Youngren W, Funderburk J. 0634 An RCT of Brief CBT for Insomnia among Primary Care Patients with Suicidal Ideation. Sleep. 2023;46(Supplement_1):A279–A.

23. Kalmbach DA, Cheng P, Ahmedani BK, Peterson EL, Reffi AN, Sagong C, et al. Cognitive-behavioral therapy for insomnia prevents and alleviates suicidal ideation: insomnia remission is a suicidolytic mechanism. Sleep. 2022;45(12).

24. Nazem S, Sun S, Barnes SM, Monteith LL, Hostetter TA, Forster JE, et al. Impact of an internet-based insomnia intervention on suicidal ideation and associated correlates in veterans at elevated suicide risk. Transl Behav Med. 2024;14(11):673–83.

25. Mournet AM, Kleiman EM. A systematic review and meta-analysis on the efficacy of sleep interventions to treat suicidal ideation. Journal of Sleep Research. 2024;33(4):e14133.

26. Moher D, Shamseer L, Clarke M, Ghersi D, Liberati A, Petticrew M, et al. Preferred reporting items for systematic review and meta-analysis protocols (PRISMA-P) 2015 statement. Systematic Reviews. 2015;4(1):1.

27. Shamseer L, Moher D, Clarke M, Ghersi D, Liberati A, Petticrew M, et al. Preferred reporting items for systematic review and meta-analysis protocols (PRISMA-P) 2015: elaboration and explanation. Bmj. 2015;350:g7647.

28. Furukawa Y, Sakata M, Yamamoto R, Nakajima S, Kikuchi S, Inoue M, et al. Components and Delivery Formats of Cognitive Behavioral Therapy for Chronic Insomnia in Adults: A Systematic Review and Component Network Meta-Analysis. JAMA Psychiatry. 2024;81(4):357–65.

29. Steinmetz L, Simon L, Feige B, Riemann D, Johann AF, Ell J, et al. Network meta-analysis examining efficacy of components of cognitive behavioural therapy for insomnia. Clin Psychol Rev. 2024;114:102507.

30. American Psychiatric Association. Diagnostic and statistical manual of mental disorders (4th ed., Text Revision). 4th edition. ed. Washington, DC;: American Psychiatric Publishing; 2000.

31. American Psychiatric Association. Diagnostic and statistical manual of mental disorders (5th ed.). 5th edition. ed. Washington, DC;: American Psychiatric Publishing; 2013.

32. American Academy of Sleep Medicine. International Classification of Sleep Disorders: American Academy of Sleep Medicine; 2014.

33. World Health Organization. ICD-10: International classification of diseases (10th revision). 2nd ed. Geneva: World Health Organization; 2004.

34. World Health Organization. ICD-11: International classification of diseases (11th revision): World Health Organization; 2022.

35. Park K, Suh S. Cognitive-Behavioral Therapy for Insomnia: A Review of the Treatment Effects on Suicide. J Sleep Med. 2017;14(2):47–54.

36. Ribeiro JD, Huang X, Fox KR, Franklin JC. Depression and hopelessness as risk factors for suicide ideation, attempts and death: meta-analysis of longitudinal studies. British Journal of Psychiatry. 2018;212(5):279–86.

37. The EndNote Team. EndNote. In: Philadelphia P, editor.: Clarivate; 2013.

38. Borenstein M, Hedges LV, Higgins JP, Rothstein HR. A basic introduction to fixed-effect and random-effects models for meta-analysis. Res Synth Methods. 2010;1(2):97–111.

39. Borenstein M, Higgins JP, Hedges LV, Rothstein HR. Basics of meta-analysis: I(2) is not an absolute measure of heterogeneity. Res Synth Methods. 2017;8(1):5–18.

40. IntHout J, Ioannidis JPA, Rovers MM, Goeman JJ. Plea for routinely presenting prediction intervals in meta-analysis. BMJ Open. 2016;6(7):e010247.

41. Mantel N, Haenszel W. Statistical aspects of the analysis of data from retrospective studies of disease. J Natl Cancer Inst. 1959;22(4):719–48.

42. Efthimiou O. Practical guide to the meta-analysis of rare events. Evidence Based Mental Health. 2018;21(2):72–6.

43. Balduzzi S, Rücker G, Schwarzer G. How to perform a meta-analysis with R: a practical tutorial. Evid Based Ment Health. 2019;22(4):153–60.

44. Sterne JA, Sutton AJ, Ioannidis JP, Terrin N, Jones DR, Lau J, et al. Recommendations for examining and interpreting funnel plot asymmetry in meta-analyses of randomised controlled trials. Bmj. 2011;343:d4002.

45. Peters JL, Sutton AJ, Jones DR, Abrams KR, Rushton L. Contour-enhanced meta-analysis funnel plots help distinguish publication bias from other causes of asymmetry. Journal of Clinical Epidemiology. 2008;61(10):991–6.

46. Sterne JAC, Savović J, Page MJ, Elbers RG, Blencowe NS, Boutron I, et al. RoB 2: a revised tool for assessing risk of bias in randomised trials. Bmj. 2019;366:4898.

47. GRADEpro GDT. GRADEpro Guideline Development Tool [Software]. McMaster University and Evidence Prime 2024 [Available from: Available from https://www.gradepro.org/.

48. Guyatt GH, Oxman AD, Vist GE, Kunz R, Falck-Ytter Y, Alonso-Coello P, et al. GRADE: an emerging consensus on rating quality of evidence and strength of recommendations. Bmj. 2008;336(7650):924–6.

49. Drake CL. The Promise of Digital CBT-I. Sleep. 2016;39(1):13–4.

