## Supplement 2 for "Cognitive behavioral therapy for insomnia as a suicide prevention strategy: a protocol for a systematic review and meta-analysis"

**Search strings**

### Ovid Medline

### 21.3.25, 58

(

Exp "suicide"/ OR

suicid*.ti,ab,kf.

)

AND

(

Exp "Sleep Wake Disorders"/ OR

Insomnia*.ti,ab,kf. OR

sleep*.ti,ab,kf.

) AND

(

Exp "Cognitive Behavioral Therapy"/ OR

CBTI.ti,ab,kf. OR

CBT.ti,ab,kf. OR

(cognitive ADJ3 (therap* OR psychotherap*)).ti,ab,kf. OR

("cognitive behavio*").ti,ab,kf. OR

(behavioral ADJ3 (therap* OR intervention* OR treatment* OR measurement*)).ti,ab,kf.

)

AND

(

Exp "Controlled Clinical Trial"/ OR

"clinical trials as topic"/ OR

"clinical trial*".ti,ab,kf. OR

random*.ti,ab,kf. OR

placebo.ti,ab,kf. OR

trial.ti.

)

### Embase

### 21.3.25, 208

(

'suicidal behavior'/exp OR

suicid*:ti,ab,kw

)

AND

(

'sleep disorder'/exp OR

Insomnia*:ti,ab,kw OR

sleep*:ti,ab,kw

)

AND

(

"Cognitive Behavioral Therapy"/exp OR

CBTI:ti,ab,kw OR

CBT:ti,ab,kw OR

(cognitive NEAR/3 (therap* OR psychotherap*)):ti,ab,kw OR

("cognitive behavio*"):ti,ab,kw OR

(behavioral NEAR/3 (therap* OR intervention* OR treatment* OR measurement*)):ti,ab,kw

)

AND

('controlled clinical trial'/exp OR 'clinical trial (topic)'/de OR 'clinical trial*':ti,ab,kw OR 'random*':ti,ab,kw OR 'placebo':ti,ab,kw OR 'trial':ti)

### Cochrane Library

### 21.3.25, 220

(

[mh "suicide"] OR

suicid*:ti,ab,kw

)

AND

(

[mh "Sleep Wake Disorders"] OR

Insomnia*:ti,ab,kw OR

sleep*:ti,ab,kw

)

AND

(

[mh "Cognitive Behavioral Therapy"] OR

CBTI OR

CBT OR

(cognitive NEAR/3 *therap*) OR

(cognitive NEXT behavio*) OR

(behavioral NEAR/3 (therap* OR intervention* OR treatment* OR measurement*))

):ti,ab,kw

### PsycInfo (EBSCO)

### 21.3.25, 32

(

suicid*

)

AND

(

Insomnia* OR

sleep*

)

AND

(

CBTI OR

CBT OR

(cognitive N2 (therap* OR psychotherap*)) OR

"cognitive behavio*" OR

(behavioral N2 (therap* OR intervention* OR treatment* OR measurement*))

)

AND

MR "clinical trial"

### ClinicalTrials.gov

### 21.3.25, 29

Condition/disease:

(suicide OR suicidal OR suicidality) AND (insomnia OR sleep)

Intervention/treatment:

(cbt OR cbti OR cognitive OR behavioral)

### Deduplication

Partially automated deduplication in EndNote moved 128 duplicates to the Trash folder.
